# Prenatal Selective Serotonin Reuptake Inhibitor Exposure, Depression and Brain Morphology in Middle Childhood: Results from the ABCD Study

**DOI:** 10.1101/2021.09.01.21262980

**Authors:** Allison L. Moreau, Michaela Voss, Isabella Hansen, Sarah E. Paul, Deanna M. Barch, Cynthia E. Rogers, Ryan Bogdan

## Abstract

**Objective:** Prenatal selective serotonin reuptake inhibitor (SSRI) exposure has been inconsistently linked to depression. Potential neural intermediaries remain understudied. We examined whether prenatal SSRI exposure is associated with depressive symptoms and brain structure during middle childhood.

**Methods:** Prenatal SSRI exposure (retrospective caregiver-report), depressive symptoms (caregiver-reported Child Behavior Checklist) and brain structure (MRI-derived subcortical volume; cortical thickness and surface area) were assessed in children (analytic ns=5,420-7,528; 235 with prenatal SSRI exposure; 9-10 years old) who completed the baseline session of the Adolescent Brain and Cognitive Development^SM^ Study. Covariates included familial (e.g., 1^st^ degree relative depression density), pregnancy (e.g., planned pregnancy), and child (e.g., birthweight) variables. Matrix spectral decomposition was used to address multiple testing.

**Results:** There was no evidence that prenatal SSRI exposure was associated with child depression after accounting for recent maternal depressive symptoms. Prenatal SSRI exposure was associated with greater left superior parietal surface area (b=145.3 mm^2^, p=0.00038) and lateral occipital cortical thickness (b=0.0272 mm, p=0.0000079), neither of which was associated with depressive symptoms.

**Conclusions:** Our findings, combined with adverse associations of prenatal exposure to maternal depression and the utility of SSRIs for treating depression, suggest that risk for child depression during middle childhood should not discourage SSRI use during pregnancy. It will be important for future work to examine associations between prenatal SSRI exposure and depression through young adulthood, when risk for depression increases.

## INTRODUCTION

Following the release of Prozac®, the first Selective Serotonin Reuptake Inhibitor (SSRI), to the United States market in 1988, antidepressant use increased by nearly 600% (from 1.8% to 10.7% of American adults)(1). As women are more likely to suffer from depression than men and the period of greatest risk occurs during reproductive years (2), it is perhaps unsurprising that among the 50% of pregnant women taking prescribed medication, SSRIs are one of the most commonly used classes of medication (3,4). However, the potential impact of prenatal SSRI exposure is poorly understood.

Prenatal SSRI exposure has been associated with a host of adverse physical outcomes (e.g., diminished fetal growth, hypertension) (5), with mixed evidence that it may (e.g., (6–12)) or may not (e.g., (13–17)) be associated with depression-related phenotypes among offspring, even when considering maternal depression (**Table 1**). Overall, larger studies have generally identified associations between prenatal SSRI exposure and depression-related outcomes, even when accounting for maternal depression and using concordant/discordant sibling designs (6,9–12), while the majority of null associations have arisen from relatively smaller samples (14,15,17). Contrasting a relatively sizable literature investigating prenatal SSRI exposure and depression-related outcomes in young children, there has been limited investigation of putative neural mechanisms through which risk may arise, despite evidence from non-human animal models that prenatal SSRI exposure may induce depression by altering brain development (e.g., (18)). Indeed, we are aware of only two structural magnetic resonance imaging (sMRI) studies of brain volume in humans, which were conducted in neonates and produced conflicting findings (19,20).^1^

**Table 1.** Summary of existing relevant literature.

| Study | Sample | Controlled for maternal depression? | Relevant Findings |
| --- | --- | --- | --- |
| **Studies Reporting That Prenatal SSRI Exposure Is Associated With Internalizing Symptoms** |  |  |  |
| Hanley et al. (2015) | Prenatal SRI exposed (n=44) and unexposed (n=66) 3-6-year-old children | Yes (during pregnancy and childhood) | Increased internalizing behaviors at age 3 and follow-up at age 6 |
| Brandlistuen et al. (2015) | Matched siblings discordant for prenatal antidepressant exposure and concordant for no exposure assessed at 18 (discordant sibling n=141, concordant sibling n=20,0039) and 36 (discordant sibling n=112, concordant sibling n=14,323) months | Yes (during pregnancy, 6 months post-partum, and lifetime history) | Increased anxiety symptoms at 36, but not 18, months after controlling for maternal depression |
| Hermansen et al. (2016) | Prenatal SSRI exposed (n=28) and unexposed (n=42 born to mothers who had unmedicated depression during pregnancy, n=33 born to healthy controls) 5-6-year-old children | Unclear, however depression symptom measures during or after pregnancy did not differ between depressed mothers taking SSRIs and unmedicated depressed mothers | Increased internalizing problems in SSRI-exposed, but not untreated depression, group, compared to healthy controls; however, the two exposure groups did not differ significantly on internalizing problems |
| Malm et al. (2016) | Prenatal SSRI exposed (n=15,729) and unexposed (n=9,651 born to mothers with unmedicated psychiatric disorders; n=7,980 whose mothers discontinued SSRI use prior to pregnancy; 31,394 born to mothers who never used SSRIs) children ages 0-14. | Indirectly in group comparisons and by excluding women using multiple psychotropic medications (which they argue is a proxy for severity) | Increased rates of depression from 12 years old in SSRI exposed group. No changes in rates of anxiety, ADHD or autism. |
| Liu et al. (2017) | Prenatal SSRI exposed (n=21,063) and unexposed (n=30,079 whose mothers discontinued SSRI use before pregnancy, 854,241 born to mothers who never used SSRIs) 0-16.5-year-old children. | Indirectly in group comparisons | Increased risk of psychiatric disorders, including mood disorders, among children exposed prenatally to antidepressants compared to children whose mothers stopped taking them before pregnancy |
| Lupattelli et al. (2018) | Prenatal SSRI exposed (n=290) and unexposed (n=3,775 born to mothers with depression prior to or during pregnancy) children followed longitudinally from 1.5-5 years of age. | Yes (during pregnancy, childhood, and lifetime history) | Late pregnancy SSRI exposure associated with increased anxious/depressed behaviors at 5 years old |
| Rommel et al. (2021) | Prenatal antidepressant exposed (n=15,892) and unexposed (n=27,096 whose mothers discontinued antidepressants before pregnancy) children ages 0-18. | Compared exposed children with those whose mothers were taking antidepressants until pregnant to control for underlying maternal psychiatric disorders | Increased risk for affective disorders for continuation group than discontinuation group (adjusted HR =1.21). However, similar increased risk for paternal antidepressant use during pregnancy suggests it may be due to parental psychopathology rather than antidepressant exposure |
| **Studies Reporting No Significant Association Between Prenatal SSRI Exposure and Internalizing Symptoms** |  |  |  |
| Nulman et al. (1997) | Prenatal SSRI exposed (n=55) and unexposed (n=84) 1.3-2.5-year-old children. | Unclear. Did not report depression levels for controls | No differences in temperament, mood or behavior problems |
| Misri et al. (2006) | Prenatal SSRI exposed (n=22) and unexposed (n=14) 4-5-year-old children. | No unmedicated depression control group, but controlled for maternal mood (during pregnancy and childhood) | No significant differences in internalizing behaviors. Maternal depressive symptoms during childhood were significantly associated with parent, but not clinician, reported internalizing symptoms. |
| Nulman et al. (2012) | Prenatal SSRI exposed (n=62), venlafaxine exposed (n=62), and unexposed (n=54 born to mothers with untreated depression; 62 born to healthy controls) 3-7-year-old children. | Yes (during pregnancy and childhood) | Antidepressant dose or duration did not predict internalizing symptoms (maternal depression during pregnancy and follow-up did) |
| Nulman et al. (2015) | Siblings (n=90), aged 3-7, discordant for prenatal SSRI exposure (n=45 exposed, 45 unexposed). | Yes (during pregnancy and childhood) | No significant differences in internalizing behaviors. Maternal depression during pregnancy and childhood was significantly associated with internalizing behaviors. |
| Grzeskowiak et al. (2016) | Prenatal antidepressant exposed (n=210) and unexposed (n=231 born to mothers with unmedicated depression, 48,737 born to healthy controls) 7-year-old children. | Yes (during pregnancy) | No significant differences in emotional symptoms after adjusting for maternal mood during pregnancy |

Given that SSRIs are one of the most commonly used forms of prescribed medication among pregnant women, and a mixed literature suggesting that SSRI use during pregnancy may be associated with depression risk among offspring, it is critical to further investigate this relationship as well as neural correlates. To this end, we used data from children (ages 9-10) who completed the baseline session of the Adolescent Brain and Cognitive Development^SM^ Study (ABCD Study^®^) to test whether prenatal SSRI exposure is associated with depressive symptoms and brain structure metrics (i.e., gray matter volume, surface area, cortical thickness) during middle childhood. Given a prior mixed literature and that children were tested during middle childhood, before heightened depression risk, we hypothesized that prenatal SSRI exposure may or may not be associated with depressive symptoms and brain structure. Secondary analyses estimated associations between child depression and brain structure as well as associations between these variables, maternal depression, and child SSRI use.

## METHODS

### Participants

Data came from children (n=11,875; M_age_ = 9.9±0.6 years; 47.85% girls; 74.13% White) born between 2005 and 2009 to 9,987 mothers through 10,801 pregnancies who completed the baseline session of the ongoing longitudinal ABCD study (data release 2.0.1; https://abcdstudy.org/; **Online Supplement**). All parents provided written informed consent after receiving a complete description of the study, and all children provided verbal assent to a research protocol approved by the institutional review board at each data collection site (n=22)^2^ throughout the United States (https://abcdstudy.org/sites/abcd-sites.html). Those with quality-controlled non-missing structural MRI and prenatal SSRI exposure data were considered for analyses (n=10,010; **Table 2**). Analytic samples ranged from 5,420-7,528 after exclusions for missingness on covariates.

**Table 2.** ABCD Sample Characteristics.

| Variable (N) | No Prenatal SSRI Exposure (N=9,730) | Prenatal SSRI Exposure |  |  | Total (N=10,010) |
| --- | --- | --- | --- | --- | --- |
|  |  | Before Maternal Knowledge of Pregnancy (N=261) | After Maternal Knowledge of Pregnancy (N=205) | Total (N=280) |  |
| Child Variables |  |  |  |  |  |
| Age in years (10,010) | 9.92 ± 0.62 | 9.94 ± 0.64 | 9.93 ± 0.63 | 9.94 ± 0.64 | 9.92 ± 0.62 |
| Sex, male (10,007) | 5,071 (52.1%) | 141 (54.0%) | 120 (58.5%) | 154 (55.0%) | 5,225 (52.2%) |
| Birthweight in oz. (9,735) | 112.61 ± 23.24 | 106.49 ± 25.75 | 105.48 ± 25.70 | 106.99 ± 25.50 | 112.45 ± 23.32 |
| Race/Ethnicity (10,010) |  |  |  |  |  |
| White | 7,249 (74.5%) | 252 (96.6%) | 201 (98.0%) | 269 (96.1%) | 7,518 (75.1%) |
| Black | 2,040 (21.0%) | 17 (6.5%) | 13 (6.3%) | 18 (6.4%) | 2,058 (20.6%) |
| Asian | 574 (5.9%) | 5 (1.9%) | 5 (2.4%) | 6 (2.1%) | 580 (5.8%) |
| Native American | 303 (3.1%) | 12 (4.6%) | 8 (3.9%) | 12 (4.3%) | 315 (3.1%) |
| Pacific Islander | 61 (0.6%) | 0 (0.0%) | 0 (0.0%) | 0 (0.0%) | 61 (0.6%) |
| Other | 659 (6.8%) | 10 (3.8%) | 6 (2.9%) | 11 (3.9%) | 670 (6.7%) |
| Hispanic | 2,071 (21.6%) | 21 (8.1%) | 14 (6.9%) | 23 (8.3%) | 2,094 (21.2%) |
| Intracranial Volume (10,005) | 1,514,365 ± 148,156.5 | 1,545,173 ± 158,780.5 | 1,556,542 ± 158,297.4 | 1,547,026 ± 157,194.8 | 1,515,279 ± 148,506.5 |
| Mean Cortical Thickness (10,005) | 2.78 ± 0.10 | 2.80 ± 0.10 | 2.80 ± 0.096 | 2.80 ± 0.10 | 2.78 ± 0.10 |
| Pregnancy and Family Variables |  |  |  |  |  |
| Planned Pregnancy (9,929) | 5,980 (62.0%) | 171 (65.8%) | 143 (70.1%) | 184 (65.9%) | 6,164 (62.1%) |
| Maternal Age at Birth (9,887) | 29.38 ± 6.20 | 31.27 ± 5.67 | 31.34 ± 5.33 | 31.27 ± 5.63 | 29.43 ± 6.19 |
| Prenatal Vitamin Use (9,834) | 9,135 (95.6%) | 248 (97.3%) | 196 (98.0%) | 267 (97.4%) | 9,402 (95.6%) |
| Household Income (9,169) |  |  |  |  |  |
| Less than \$50k | 2,661 (29.9%) | 43 (17.4%) | 30 (15.7%) | 46 (17.4%) | 2,707 (29.5%) |
| \$50k-\$74,999 | 1,222 (13.7%) | 40 (16.2%) | 28 (14.7%) | 42 (15.9%) | 1,264 (13.8%) |
| \$75k-\$99,999 | 1,288 (14.5%) | 42 (17.0%) | 30 (15.7%) | 45 (17.0%) | 1,333 (14.5%) |
| \$100k-\$199,999 | 2,706 (30.4%) | 93 (37.7%) | 73 (38.2%) | 97 (36.7%) | 2,803 (30.6%) |
| \$200k or more | 1,028 (11.5%) | 29 (11.7%) | 30 (15.7%) | 34 (12.9%) | 1,062 (11.6%) |
| Maternal Education: Years (9,656) | 15.18 ± 2.61 | 16.04 ± 2.01 | 15.97 ± 2.00 | 16.04 ± 2.01 | 15.20 ± 2.60 |
| Maternal Dep History (9,651) | 1,983 (21.2%) | 170 (65.6%) | 124 (60.5%) | 180 (64.7%) | 2,163 (22.4%) |
| Maternal Recent Dep Sxs (8,842) | 53.74 ± 5.70 | 59.17 ± 7.67 | 58.87 ± 7.65 | 58.98 ± 7.63 | 53.89 ± 5.83 |
| Family Dep History, % (9,253) | 11 ± 15 | 24 ± 20 | 23 ± 19 | 24 ± 20 | 11 ± 16 |
| Twin/triplet in study (10,010) | 1,789 (18.4%) | 63 (24.1%) | 51 (24.9%) | 64 (22.9%) | 1,853 (18.5%) |
| Prenatal Substance Exposure Before Knowing of Pregnancy |  |  |  |  |  |
| Tobacco before (9,979) | 1,227 (12.6%) | 34 (13.1%) | 19 (9.3%) | 36 (12.9%) | 1,263 (12.7%) |
| Alcohol before (9,690) | 2,281 (24.2%) | 95 (37.0%) | 69 (34.2%) | 101 (36.7%) | 2,382 (24.6%) |
| Cannabis before (9,945) | 492 (5.1%) | 16 (6.2%) | 8 (3.9%) | 18 (6.5%) | 510 (5.1%) |
| *Prenatal Substance Exposure After Knowing of Pregnancy* |  |  |  |  |  |
| Tobacco after (9,997) | 428 (4.4%) | 11 (4.2%) | 6 (2.9%) | 11 (3.9%) | 439 (4.4%) |
| Alcohol after (9,993) | 216 (2.2%) | 8 (3.1%) | 8 (3.9%) | 8 (2.9%) | 224 (2.2%) |
| Cannabis after (9,995) | 156 (1.6%) | 5 (1.9%) | 3 (1.5%) | 6 (2.1%) | 162 (1.6%) |

Notes: Dep = Depression; Sxs = Symptoms; SSRI = Selective Serotonin Reuptake Inhibitor; Oz = ounces. Values are either N (%) or mean ± standard deviation.

### Measures

Data for all investigated variables are displayed in **Table 2**.

### Prenatal Exposure to SSRIs and Other Medications

A parent or caregiver (89.1% biological mothers) retrospectively reported medications used by the mother during pregnancy, before and after maternal knowledge of pregnancy. Two-hundred and eighty (2.80%) children were reported to be prenatally exposed to SSRIs at some point during pregnancy, with 261 exposed prior to and 205 exposed following maternal knowledge of pregnancy. The majority of individuals were exposed both prior to and after maternal knowledge of pregnancy (n=186); 75 were exposed only prior to maternal knowledge, and 19 were exposed only after maternal knowledge of pregnancy.

### Child Depressive Symptoms

The Depression DSM-5 Scale of the Child Behavior Checklist (CBCL)(21) was used to assess child depressive symptoms within the past six months according to parent/caregiver report.

### Child Brain Structure

FreeSurfer v5.3 (http://surfer.nmr.mgh.harvard.edu/) was used to estimate the following global and regional structural MRI metrics: *global:* total brain volume, mean cortical thickness, total cortical surface area; *regional:* Desikan-Killiany cortical parcellation regions (22) (n=35/hemisphere, including hemisphere total) for cortical thickness and surface area, as well as segmented subcortical volumes (23) (n=31, including left and right hemisphere cortical totals). Acquisition and processing for the ABCD study are described in the **Online Supplement** and (24) and (25), respectively.

### SSRI Exposure During Childhood

Caregivers reported whether the child took any prescribed medications within the past two weeks; 140 children (1.4%) were reported to have used SSRIs.

### Maternal Depression

The parent/caregiver who completed the baseline questionnaires reported whether the child’s biological mother had ever experienced depression. Caregivers also completed the Adult Self-Report (ASR) questionnaire, which assesses their own past-six-month psychopathology, including depression; we used self-reported depression data from biological mothers only (n=8,842).

### Covariates

The following variables were considered as covariates in analyses: reported child sex, race [i.e., White, Black, Asian, Native Hawaiian or Pacific Islander, Native American or Alaskan, Other, Mixed Race], ethnicity (Hispanic/Latinx), age, and birthweight, as well as household income, maternal education, whether the pregnancy was planned, maternal age at birth, maternal lifetime history of depression, percentage of family members (biological parents, grandparents, siblings, aunts, and uncles) with a history of depression, prenatal exposure to prenatal vitamins, tobacco, marijuana, and alcohol and whether the child had a reported twin or triplet in the study (**Online Supplement**). For all imaging analyses, scanner ID was used instead of site ID as a grouping variable. Regional volume and surface area analyses also included intracranial volume (ICV) to account for total brain volume while regional cortical thickness analyses included average whole-brain cortical thickness (**Online Supplement**).

### Analyses

As the sample contains twin and non-twin siblings, as well as 22 research sites, linear mixed effect models were used to nest data by family and site/scanner separately using the lme4 (version 1.1-21) package in R (version 3.6.0). All analyses accounted for the fixed effect covariates described above.

#### Primary Analyses: Prenatal SSRI Exposure

First, we estimated associations between prenatal SSRI exposure at any point during pregnancy and depressive symptoms in children. Second, we estimated whether prenatal SSRI exposure was associated with brain structure using a two-tiered analytic approach. Here, we *first* tested *a priori* regions of interest (ROIs) previously associated with adolescent and adult depression in the largest available meta analyses^3^ (26–28) (n=42)^4^: hippocampus and amygdala volume; *surface area* in lingual, superior frontal, postcentral, pericalcarine, lateral occipital, medial orbitofrontal and precentral gyri and left and right hemispheres overall; *cortical thickness* in medial orbitofrontal cortex, fusiform, insula, rostral anterior, caudal anterior and posterior cingulate cortex, middle and inferior temporal, supramarginal, inferior frontal (pars opercularis) and lingual gyri. Multiple testing was corrected for using Matrix Spectral Decomposition (MatSpD), which accounts for the correlation between brain structure metrics to estimate the number of independent tests conducted (25 independent tests estimated; adjusted Bonferroni alpha level of 0.00205). *Then*, we conducted exploratory SSRI-brain structure analyses of all remaining 132 neural regional phenotypes available. MatSpD estimated 91.4 independent tests from 174 total tests (including *a priori* tests above) resulting in an adjusted Bonferroni alpha level of 0.000561 for exploratory analyses.

Any significant associations were subsequently examined when stratified according to the timing of prenatal SSRI exposure (before/after maternal knowledge of pregnancy) and when controlling for recent maternal depressive symptoms. In addition, significant associations were analyzed when excluding children who were currently taking SSRIs, exposed to illicit substances prenatally, born at extreme levels of prematurity, or whose biological mothers were not the parent/caregiver respondent (**Online Supplement**). To account for potential confounding effects of genomic liability not accounted for by familial density of depression and maternal lifetime depression, we examined significant associations when accounting for child polygenic risk scores (PRS) for depression (**Online Supplement**). Finally, to evaluate the specificity of observed findings, we tested whether prenatal exposure to the antidepressant Wellbutrin (n=63), antihistamines (n=78), or prescription opioid pain medications (n=100) were associated with variability in phenotypes linked to prenatal SSRI exposure (**Online Supplement**).

#### Secondary Analyses: Child Brain Structure Correlates of Child and Maternal Depression and Child SSRI Use

Secondary analyses of associations between child depression and brain structure and their relation to lifetime maternal depression, recent maternal depression, and child SSRI use were conducted (**Online Supplement)**.

## RESULTS

### Prenatal SSRI exposure and child depression outcomes

Prenatal SSRI exposure (n=235)^5^ was associated with a small increase in depressive symptoms among children, after accounting for covariates previously described (b=1.18 [0.44, 1.93], t=3.11, ΔR^2^=0.0013, p=0.0019; **Figure 1**). *Post hoc* regressions revealed that this association: A) was primarily driven by prenatal SSRI exposure before (n=219; b=1.16, ΔR^2^=0.0012, p=0.0032), but not after (n=172; b=0.84, ΔR^2^=0.00054, p=0.056), maternal knowledge of pregnancy, B) remained when excluding children currently taking SSRIs, exposed prenatally to illicit substances, born very prematurely, or whose biological mother was not the caregiver respondent, C) was largely unaltered in effect size when accounting for child polygenic risk for depression, though no longer significant due to sample size reductions, and D) was no longer significant when also controlling for recent maternal depressive symptoms (b=0.179, p=0.63) (**Online Supplement**).^6^

**Figure 1.**
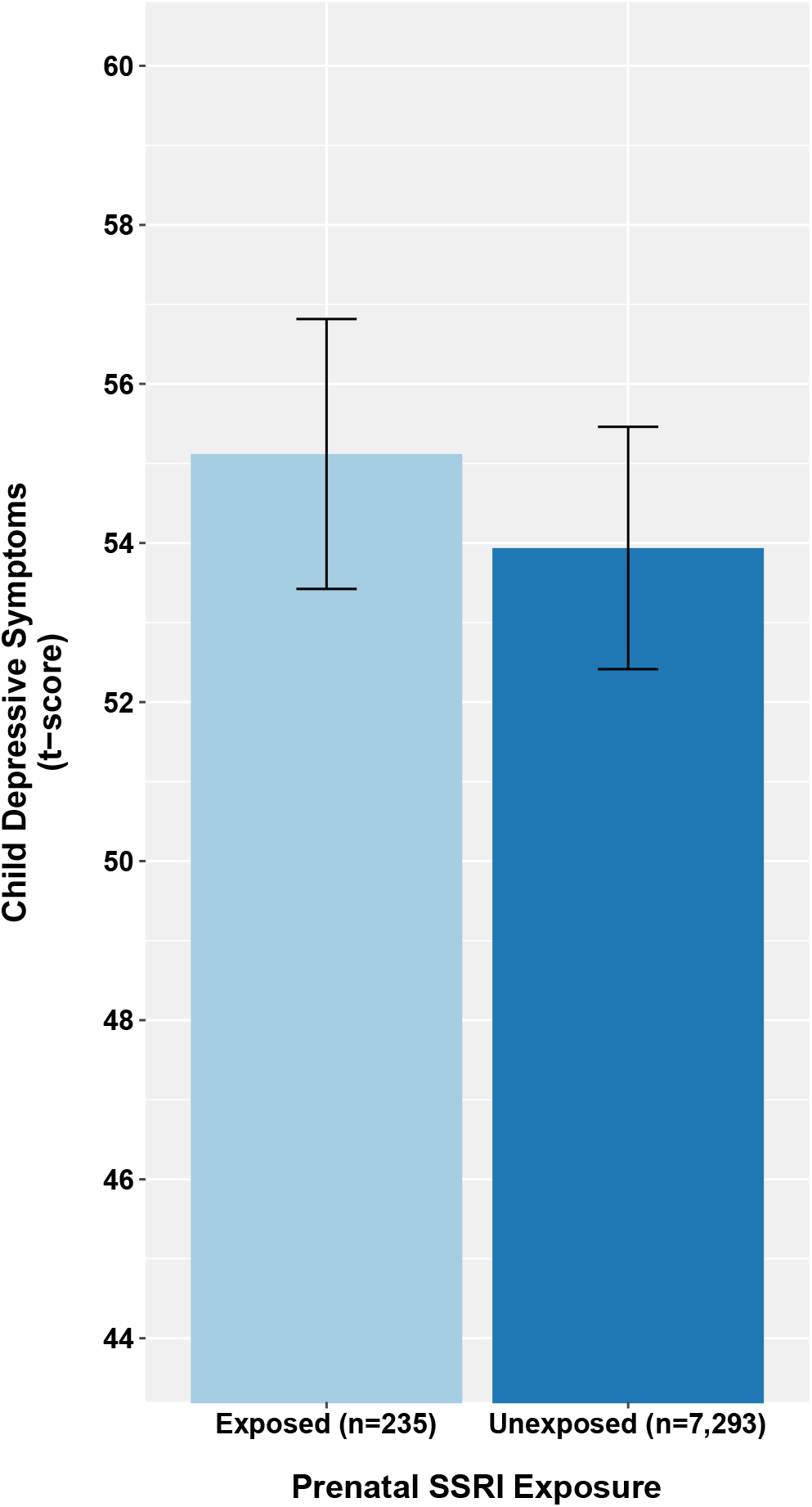
Children Prenatally Exposed to SSRIs have Elevated Depressive Symptoms. <u>Notes:</u> Depression symptom measure: Child Behavior Checklist. Error bars represent 95% confidence intervals. Ns represent the analytic sample included in the regression analysis after listwise deletion for missing covariate data. Estimated marginal means were extracted from mixed effects models that control for all covariates, including lifetime history of maternal depression, and the dependency in the data.

### Prenatal SSRI exposure and child brain outcomes

Nominally significant findings are reported in the **Online Supplement**.

#### Analyses of a priori depression ROIs

Prenatal SSRI exposure was not significantly associated with any *a priori* brain structure phenotypes after correction for multiple testing (all p’s>0.002; **Table S1**).

#### Exploratory analyses in all brain ROIs

Prenatal SSRI exposure was associated with thicker left lateral occipital cortex (b=0.0272 mm [0.0153, 0.0392], t=4.47, ΔR^2^=0.0011, p=0.0000079; **Figure 2a**; **Table S1**) and larger left superior parietal surface area (b=145.3 mm^2^ [65.4, 225.2], t=3.55, ΔR^2^=0.00095, p=0.00038; **Figure 2b**; **Table S1**) after correction for multiple testing. These findings remained significant both before and after maternal knowledge of pregnancy and when accounting for recent maternal depressive symptoms and child depression PRS, as well as when excluding children currently taking SSRIs, exposed prenatally to illicit substances, born very prematurely, or whose biological mother was not the caregiver respondent (**Online Supplement**).

**Figure 2.**
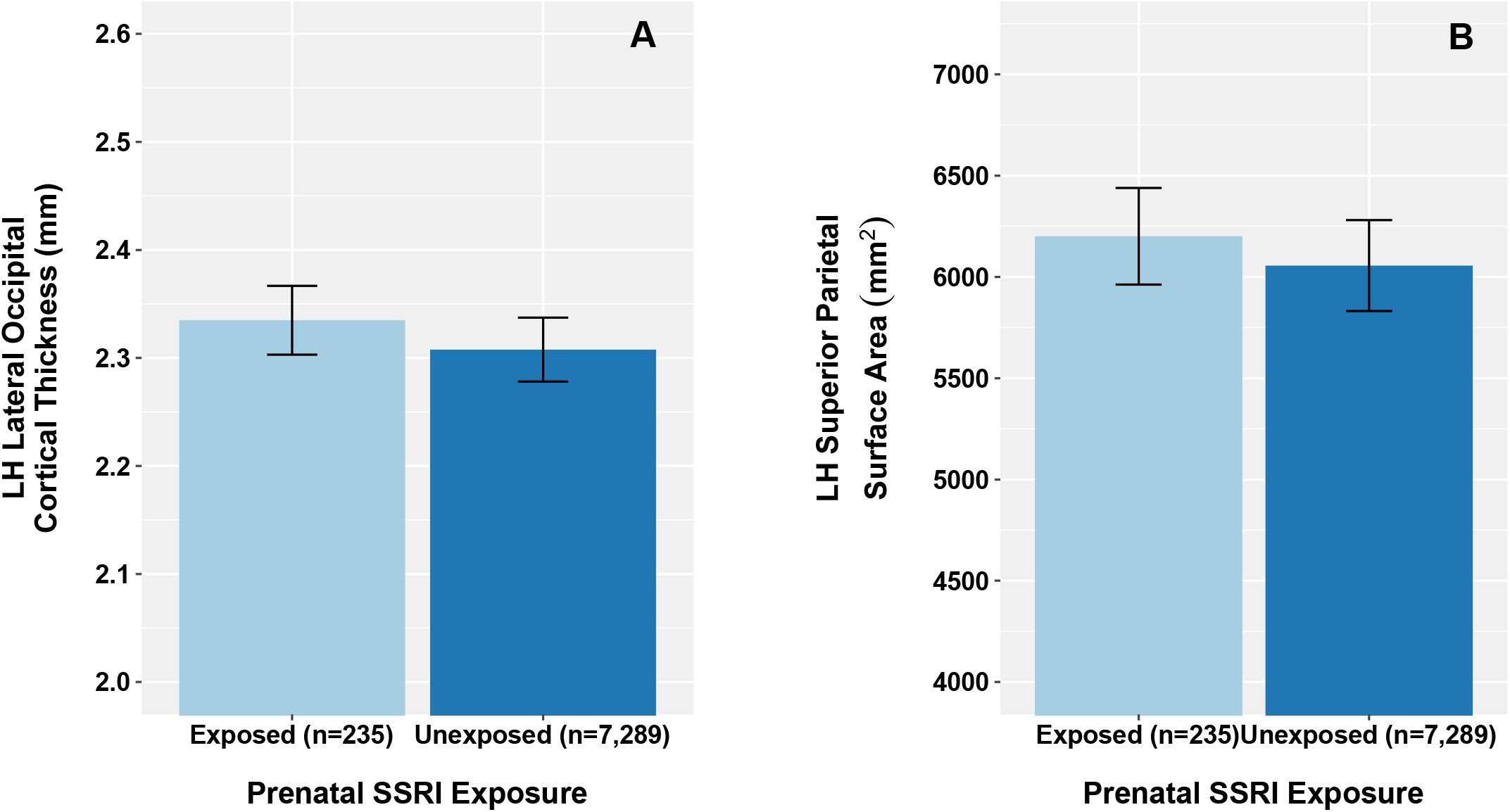
Children Prenatally Exposed to SSRIs Have Thicker Left Lateral Occipital Cortex and Larger Left Superior Parietal Surface Area. <u>Notes</u>: Error bars represent 95% confidence intervals. Ns represent the analytic sample included in the regression analysis after listwise deletion for missing covariate data. Estimated marginal means were extracted from mixed effects models that control for all covariates, including lifetime history of maternal depression, and the dependency in the data.

### Prenatal exposure to other medications and child brain and depression outcomes

Prenatal exposure to Wellbutrin (n=63), antihistamines (n=78), and prescription pain medication (n=100) were not significantly associated with child depressive symptoms or brain structure (**Online Supplement**).

### Secondary Analyses

All nominally significant findings are described in **Supplemental Results**.

#### Maternal depression and child depression and brain structure

Lifetime maternal depression and recent maternal depressive symptoms were both associated with increased child depressive symptoms (*lifetime*: b=1.22 [0.841, 1.61], t=6.24, ΔR^2^=0.0045, p=4.58e-10; *recent*: b=0.348 [0.325, 0.371], t=29.8, ΔR^2^=0.11, p=5.45e-182; **Figure S1**), but not child brain structure after correcting for multiple testing (all p’s >0.002; **Tables S2 and S3**).

#### Child brain structure and depressive symptoms

Smaller total bilateral surface area was associated with more child depressive symptoms (*total*: b=-0.0000168, ΔR^2^=0.0017, p=0.000115; *left*: b=-0.0000320, ΔR^2^=0.0015, p=0.000236; *right:* b=-0.0000345, ΔR^2^=0.0018, p=0.0000612; **Figure S2**). Exploratory analyses revealed that child depressive symptoms were also significantly associated with total brain volume (b=-0.00000270, ΔR^2^=0.0023, p=0.000115) as well as left and right hemisphere cortical volume (*left*: b=- 0.0000109, ΔR^2^=0.0018, p=0.0000622; *right*: b=-0.0000112, ΔR^2^=0.0019, p=0.0000409; **Figure S2; Table S4**).

#### Child SSRI Use

Child SSRI use was associated with higher child depressive symptoms (b=7.10 [6.05, 8.08], t=13.6, ΔR^2^=0.031, p<2e-16) and larger surface area in the right isthmus cingulate (b=51.9 mm^2^ [23.9, 79.8], t=3.62, ΔR^2^=0.0037, p=0.000294; **Table S5; Figure S3**). Maternal depression (recent and lifetime history) was not significantly associated with child SSRI use (*recent*: odds ratio=1.056, z=1.525, p=0.127; *lifetime*: odds ratio=1.747, z=1.70, p=0.090).

## DISCUSSION

Two primary findings emerged from our investigation of retrospectively reported prenatal SSRI exposure, depression, and brain structure during middle childhood. *First*, we found that prenatal SSRI exposure was not associated with depression symptoms during middle childhood independent of recent maternal depressive symptoms. *Second*, prenatal SSRI exposure was associated with larger surface area in the left superior parietal region and thicker left lateral occipital cortex. However, this variability in brain structure was not associated with child depressive symptoms and effect sizes were small. Given risks associated with prenatal exposure to maternal depression (29), the effectiveness of treating depression during pregnancy with SSRIs (30), and the stronger associations between maternal depression and child depression, our data suggest that concern for depression risk following SSRI exposure should not discourage SSRI use during pregnancy. It will be important for future work to extend this investigation into later adolescence and young adulthood when depression risk markedly increases.

### Prenatal SSRI Exposure and Depressive Symptoms During Middle Childhood

The initial link we observed between prenatal SSRI exposure and a small elevation in depressive symptoms in children was robust to the inclusion of lifetime history of maternal depression and depression density among first-degree relatives, but not recent (i.e., past-six-months) maternal depressive symptoms. Our observations contrast with large studies showing that prenatal SSRI exposure is associated with elevated risk for depression or internalizing symptoms (**Table 1**; (6– 12)). Notably, over half of these studies did not explicitly model any form of maternal depression in analyses (8–10,12) and only a few (7,11) evaluated recent maternal depressive symptoms, though the children in these studies were younger than the current one. Along these lines, our observations mirror prior null associations observed for child internalizing symptoms at younger ages in studies that accounted for maternal depression during pregnancy and childhood ((13–17)). However, the vast majority of these studies would be underpowered to detect the effect size we observed here, prior to the inclusion of recent maternal depressive symptoms. That recent maternal depressive symptoms, but not lifetime history of maternal depression and familial density of depression, accounted for the significant association between prenatal SSRI exposure and child depressive symptoms highlights potential depressogenic effects of postnatal rearing environments associated with maternal depression, which has been well-documented in the literature (31). Indeed, recent maternal depressive symptoms explained 11% of the variance in child depressive symptoms, while prenatal SSRI exposure explained only 0.13%. However, it is also possible that the association between recent maternal depression symptoms and maternal-rated child depression is inflated due to increased negative perceptions among depressed individuals (32). Nonetheless, it will be important for future studies of prenatal SSRI exposure to include maternal depression in analytic models.

Experimental non-human animal studies have found evidence that prenatal SSRI exposure is depressogenic (33). However, our associations, before controlling for recent maternal depression, are discrepant with regard to timing of exposure and offspring assessment in these models. For example, one study of mice found that exposure during the equivalent period of the third trimester in humans was associated with depressive-like behaviors during late adolescence/young adulthood (33). Our observed association was restricted to exposure prior to maternal knowledge of pregnancy, which occurred on average well before the third trimester (6.8 weeks). As non-human animal models and some human studies (e.g., (9)) suggest that the depressogenic effects of prenatal SSRI exposure do not emerge until later adolescence, it remains plausible that effects will be larger and observed for different timings of exposure at later developmental stages (e.g., late adolescence, young adulthood), when depression risk rises.

Given that prenatal SSRI exposure was not independently associated with child depressive symptoms, when considering recent maternal depression, our findings suggest that effectively treating maternal depression is crucial to minimize adverse effects on children. SSRIs and other antidepressants are first-line treatments for depression that can not only improve maternal depression, but possibly also that of the mother’s children as well (34). Empirically-supported psychotherapies represent another first-line treatment option for pregnant women and mothers (35). Digital administration of psychotherapy, which was recently shown to reduce depression, anxiety, and stress during pregnancy and the postpartum period in a meta-analysis of randomized controlled trials, may increase accessibility and reduce barriers (36).

### Prenatal SSRI Exposure and Brain Structure during Middle Childhood

If prenatal SSRI exposure influences child behavior, it would presumably do so by influencing the brain. For instance, prenatal SSRIs lead to decreases in S100B, a protein that mediates neonatal neuronal outgrowth and survival (37). However, we found limited evidence that prenatal SSRI exposure is associated with gray matter-related brain structure (i.e., volume, cortical thickness, surface area) during middle childhood. Indeed, only left superior parietal surface area and left lateral occipital cortical thickness were significantly associated with prenatal SSRI exposure after accounting for multiple testing; these effects were small (both ∼0.1%) and in the direction of increased size, which would not align with the S100B hypothesis. Additionally, these regional metrics have not been significantly associated with Major Depressive Disorder among adolescents or adults in recent large meta-analyses (26,28). Of note, however, an infant study also found increased brain structure (i.e., volume) in the left occipital cortex of infants prenatally exposed to SSRIs (20). In the present study, prenatal SSRI exposure was not associated with child depressive symptoms, independent of recent maternal depressive symptoms, precluding an interpretation of any mediating or moderating effects of brain structure. Still, it remains possible that the small brain structure differences we observed may be associated with subsequent depression, or that SSRI exposure and its correlates are associated with regional brain structure during earlier and/or later development.

### Secondary Analyses: Brain Structure, Depressive Symptoms, and SSRI Use Among Children

Depressive symptoms during middle childhood were associated with smaller global brain structure (i.e., total surface area and brain volume). Much like the other associations we observed, these effects were small, accounting for <0.25% of variance in depressive symptoms. Inconsistent with prior studies (26–28), we found no evidence of regional associations; this divergence may be attributable to the relatively younger age of our sample (but see also, e.g., (38)). Although maternal depression history and child SSRI use were unsurprisingly associated with greater depressive symptoms in children, only child SSRI use was associated with brain structure. Children taking SSRIs had increased right isthmus cingulate surface area. Interestingly, maternal depression (lifetime or recent) was not significantly associated with child SSRI use.

### Limitations

Some study limitations are noteworthy. *First*, we relied on ∼10 year retrospective report of SSRI use during pregnancy, which may result in misclassification (39). In the ABCD Study, 2.8% of children had mothers who used SSRIs during pregnancy, which is on the lower end of prevalence estimates in the United States during their time of birth, which range from 2.6-15% (3,4). This may reflect an underestimate due to retrospective recall and/or ascertainment bias associated with participating in a study of child health. *Second*, our study is cross sectional and does not follow children into peak depression risk (i.e., later adolescence-young adulthood; (40)). As such, it remains possible that child depression and brain structure or longitudinal trajectories of these variables are associated with prenatal SSRI exposure. As the ABCD study will follow recruited children through adolescence and into young adulthood, future studies will have the opportunity to conduct longitudinal analyses as children enter the period of developmental risk for depression. *Third*, although this represents the largest study of prenatal SSRI exposure, brain structure and depression during middle childhood to our knowledge, our power was limited by a proportionally small number of participants exposed to SSRIs prenatally, especially for analyses of exposure timing (i.e., before or after maternal knowledge of pregnancy); this is likely compounded by unmeasured heterogeneity (e.g., duration) in exposure.

*Fourth*, despite modeling many familial, pregnancy, and child-related variables that may potentially confound associations, the potential role of unmeasured confounds or alternative derivations cannot be discounted. For example, only 180 of the 280 mothers who took SSRIs during their pregnancy had a reported lifetime history of depression, so it is possible that other medical conditions not accounted for in our models may impact results. Further, while we included lifetime maternal depression, 1^st^ degree relative lifetime depression density, child polygenic risk for depression, and recent maternal depressive symptoms in our analyses, we were unable to estimate potential associations between maternal depression during pregnancy and child outcomes in our models as these data were not collected. Some prior studies have found that perinatal depression, but not prenatal SSRI exposure, is associated with childhood internalizing symptoms (15,16). *Fifth*, as is standard practice for this age of children, we relied on caregiver report of child depression and there is some evidence that unlike externalizing symptoms, internalizing symptoms may not be well captured by observer report (41). *Sixth*, the analyzed dataset contained limited data on the timing of antidepressant exposure (i.e., pre and post maternal knowledge of pregnancy), which limits our ability to examine time-dependent associations (11).

## Conclusions and Clinical Implications

These data suggest that potential risk for depression during middle childhood should not dissuade the use of SSRIs during pregnancy. Specifically, our observed associations were small in magnitude, and the link between prenatal SSRI exposure and child depressive symptoms was not independent of recent maternal depressive symptoms. Considering adverse offspring outcomes associated with maternal depression during pregnancy (42) and increased risk of relapse following SSRI discontinuation during pregnancy (30), these findings assuage concerns regarding depression risk among offspring prenatally exposed to SSRIs. It will be important for future work to follow children into peak periods of risk to further evaluate potential associations between prenatal SSRI exposure, brain structure, and depression.

## Data Availability

The data in these analyses came from the publicly available Adolescent Brain and Cognitive Development (ABCD) Study dataset (release version 2.0.1) housed on the National Institute of Mental Health's Data Archive (NDA).

https://nda.nih.gov/

## Acknowledgements

Data for this study were provided by the Adolescent Brain Cognitive Development Study℠ (ABCD Study^®^), which was funded by awards U01DA041022, U01DA041025, U01DA041028, U01DA041048, U01DA041089, U01DA041093, U01DA041106, U01DA041117, U01DA041120, U01DA041134, U01DA041148, U01DA041156, U01DA041174, U24DA041123, and U24DA041147 from the NIH and additional federal partners (https://abcdstudy.org/federal-partners.html). Authors received funding support from NIH: Dr. Rogers (R01-DA046224, R34-DA050272, R01-MH121877, R01-MH113570), Dr. Barch (R01-MH113883, R01-MH066031, U01-MH109589, U01-A005020803, R01-MH090786), Dr. Bogdan (R01-AG045231, R01-HD083614, R01-AG052564, R21-AA027827, R01-DA046224). The sponsors had no role in the design and conduct of the study; collection, management, analysis, and interpretation of the data; preparation, review, or approval of the manuscript; and decision to submit the manuscript for publication.

## Footnotes

1 One study (19) found no differences in global or regional brain volume between SSRI-exposed and unexposed (healthy control or untreated prenatal maternal depression) neonates (total N=204), while the other (20) found that SSRI-exposed neonates are characterized by larger gray matter volumes in the left occipital gyrus and right amygdala, insula, and superior frontal relative to healthy controls and infants exposed to untreated maternal depression (total N=98). Three additional diffusion tensor imaging (DTI) studies have been conducted which yielded equivocal results. Two identified lower fractional anisotropy (FA) in several regions of prenatally exposed infants (19,43); one of these also found higher FA in other regions (43). The third found increased white matter connectivity using a different approach (20) (for review, see (44)).

2 Cornell University was an original collection site that collected data from 34 participants, before being moved to Yale University. ABCD documentation reports 21 data collection sites and does not list Cornell; our analyses nested data based on 22 data collection sites, including the original Cornell site.

3 The adolescent/adult depression literature was relied upon due to the limited available literature on SSRI exposure and brain structure as well as childhood depression.

4 Regions were explored bilaterally in our analyses, regardless of whether the original finding was lateralized. Due to the important role of the amygdala in processing emotionally salient stimuli as well as associations with SSRI exposure, we included this as an *a priori* ROI despite its trending association in a meta-analysis of depression (p=0.09).

5 While the overall sample with non-missing prenatal medication and MRI data (n=10,010) contained 280 children with prenatal SSRI exposure before or after maternal knowledge of pregnancy, the analytic sample used in this regression after listwise deletion of subjects with missing data contained 235 children with prenatal SSRI exposure before or after maternal knowledge of pregnancy.

6 Our consideration of recent maternal depressive symptoms was limited to children whose biological mother brought them in for the study visit (n=8,842 out of 10,010; analytic n following listwise deletion = 6,946); nonetheless, this reduction in sample size alone was not responsible for attenuating the association between child depression and prenatal SSRI exposure as restricting our analyses to this subset but not including recent maternal depression symptoms resulted in significant associations between prenatal SSRI exposure and child depression (b=1.16, p=0.0035). Further, the percentage of the analytic sample exposed prenatally to SSRIs was nearly identical in the original and post hoc analysis (3.1 and 3.2%, respectively).

